# BNT162b2 post-exposure-prophylaxis against COVID-19

**DOI:** 10.1101/2022.01.07.22268869

**Authors:** Zohar Shmuelian, Yehuda Warszawer, Omri Or, Sagit Arbel-Alon, Hilla Giladi, Meytal Avgil Tsadok, Roy Cohen, Galit Shefer, Antonello Maruotti, Giovanna Jona Lasinio, Eithan Galun

## Abstract

**Background:** During the COVID-19 pandemic, post-exposure-prophylaxis is not a practice. Following exposure, only patient isolation is imposed. Moreover, no therapeutic prevention approach is applied. We asked whether evidence exists for reduced mortality rate following post-exposure-prophylaxis.

**Methods:** To estimate the effectiveness of post-exposure-prophylaxis, we obtained data from the Israeli Ministry of Health (MoH) registry. The study population consisted of Israeli residents aged 12 years and older, identified for the first time as PCR-positive for SARS-CoV-2, between December 20^th^, 2020 (the beginning of the vaccination campaign) and October 7^th^, 2021. We compared “recently injected” patients - that proved PCR-positive on the same day or on one of the five consecutive days after first vaccination (representing an unintended post-exposure-prophylaxis), to unvaccinated control group.

**Results:** Among Israeli residents identified PCR-positive for SARS-CoV-2, 11,690 were found positive on the day they received their first vaccine injection (BNT162b2) or on one of the 5 days thereafter. In patients over 65 years, 143 deaths occurred among 1413 recently injected (10.12%) compared to 280 deaths among the 1413 unvaccinated (19.82%), odd ratio (OR) 0.46 (95% confidence interval (CI), 0.36 to 0.57; P<0.001). The most significant reduction in the death toll was observed among the 55 to 64 age group, with 8 deaths occurring among the 1322 recently injected (0.61%) compared to 43 deaths among the 1322 unvaccinated control (3.25%), OR 0.18 (95% CI, 0.07 to 0.39; P<0.001).

**Conclusion:** Post-exposure-prophylaxis is effective against death in COVID-19 infection.

**Israeli MoH Registry Number:** HMO-0372-20

## Introduction

SARS-CoV-2 vaccination has proved effective in preventing clinical COVID-19 infection (1). Numerous drugs had shown some benefit to patients with COVID-19. These include the following compounds: Remdesivir (2), Paxlovid which received an emergency use in PCR-positive patients (3), the anti-inflammatory drugs Dexamethasone (4) and anti-IL6 receptor (5) and some compounds in clinical development such as Molnupiravir (6). However, the therapeutic benefit from most of these treatments is still suboptimal so additional therapeutic approaches are needed to reduce the loss of human lives. One potential clinical approach is post exposure vaccination prophylaxis. Post exposure vaccination prophylaxis is an old approach used to attenuate severe infections by busting an efficient immune response. This is adopted following bacterial infections in the case of tetanus, but is more common following exposure to viral infections. Although the post exposure approach is many times specific to the viral infection context, as for hepatitis B virus (7) or after a bite from a rabies infected dog, the concept is generally the same (8). Thus, active vaccination with the attenuated/killed pathogen or viral associated protein is a very effective mean to attenuate and almost eliminate any infection related symptoms. This approach is now also suggested for Ebola virus infection (9). In accordance, it was recently shown that the overall neutralizing potency of plasma is greater following vaccination compared to natural infection with SARS-CoV-2 (10). In the case of vaccinating a patient with COVID-19 respiratory illness, it is expected that upon vaccination an efficient immune response against SARS-CoV-2 will develop in a remote site from the pneumonitis developed in the lung, possibly preventing respiratory deterioration. Post exposure vaccination prophylaxis is particular relevant in light of the waning immunity against COVID-19 after BNT162b2 vaccination (11-14). We wish to determine in the real world, whether vaccinating patients who were already infected by SARS-CoV-2 will benefit from post-exposure prophylaxis vaccination. This could be in particular relevant as a measure undertaken together with isolation strategies following exposure. We performed this investigation by a data mining study on an available national database.

## Methods

### Study design

In this observational study, we analyzed nationwide surveillance data obtained from the Israeli Ministry of Health (MoH) registry to assess the effectiveness of the BNT162b2 vaccine as post exposure prophylaxis approach against the clinical outcomes of SARS-CoV-2 infection. Our study population consisted of Israeli residents aged 12 years and older, identified for the first time as PCR-positive for SARS-CoV-2, between December 20^th^, 2020 (the beginning of the vaccination campaign) and October 7^th^, 2021, and did not include patients that were found PCR-positive 6 days or later after they received the first vaccine injection (Fig. S1). The data on each patient included: age, gender, first positive PCR date, first vaccination date, hospitalization dates (arrival and discharged) and date of death. We compared two groups: Group1) the “**recently injected**” test group - patients proved PCR-positive on the same day or on one of the five consecutive days after receiving their first vaccine injection. This group was chosen because it practically represents an unintended post exposure prophylaxis treatment. Group2) The control **unvaccinated** group. To establish this control group, we matched the “recently injected” group to the unvaccinated controls using the following variables: gender, age and date of the first positive PCR, variables associated with the severity of COVID-19.

Anonymized data, without any personal identifiers, were used in this analysis. This study was approved by the Hadassah Hebrew University Hospital institutional review board (IRB approval number HMO-0372-20). The study was exempt from the requirement for informed consent.

### Outcomes

The primary outcome was COVID-19 related death (reported to the Israeli MOH) within 60 days after the first positive PCR date. Secondary outcomes were: 1) Hospitalization for COVID-19 (hospitalization in COVID-19 designated department, at the time interval between 2 days before PCR-positive and 21 days thereafter) Fig. S2; 2) Hospitalization duration.

### Statistical Analysis

For estimating the treatment effect in this observational study and to produce inferences that are more robust and less sensitive to modelling assumptions, we perform propensity score matching (15) so that we can further control for measured confounding variables, namely the gender, age and the date of the first positive PCR. One by one, each treated unit is paired with an available control unit that has the closest propensity score to it, the so called 1:1 nearest neighbor matching. Any remaining control units are left unmatched and excluded from further analysis. The goal of matching is to produce covariate balance, that is, for the distributions of covariates in the two groups to be approximately equal to each other, as they would be in a successful randomized experiment. The importance of covariate balance is that it allows for increased robustness to the choice of model used to estimate the treatment effect; in perfectly balanced samples, a simple difference in means can be a valid treatment effect estimate (loveplot: https://ngreifer.github.io/cobalt/reference/love.plot.html). Survival curves for the recently injected and unvaccinated groups were estimated using the Kaplan–Meier estimator (16, 17) analyzing death, and hospitalization probability. Fisher exact test was used to assess the association between vaccination status and the main outcomes distinguishing by age group and gender. For significant association at Fisher exact test, we calculated odds ratio (OR) and 95% confidence interval (CI). A negative binomial generalized linear model was estimated to evaluate the relation between duration of hospitalization and all interactions among age groups, vaccination status and gender. Count data may be distributed as Negative Binomial if the rate, at which events occur, is heterogeneous, and consequently the counts are characterized by overdispersion compared to the Poisson (as typically happens in our length of hospitalization data). The Poisson distribution is nested within the Negative Binomial, in the sense that if no overdispersion/heterogeneity is present, the Negative Binomial distribution converges to the Poisson distribution.

## Results

### Study population

Our study population included 271,710 Israeli residents that were identified PCR-positive for SARS-CoV-2 during the period between December 20^th^, 2020 and October 7^th^, 2021, (Fig.1). Among them, 11,690 were found positive on the same day they received their first vaccine injection (BNT162b2, Pfizer, USA) or on one of the 5 days thereafter i.e., “recently injected” group. We matched the recently injected group to unvaccinated controls according to the following variables: age, age group, gender and date (Fig.1). The characteristics of the matched unvaccinated control and the recently injected group are shown in Table 1. Notably, there are no differences in demographic characteristics between the two groups (Fig. S3).

**Table 1.**
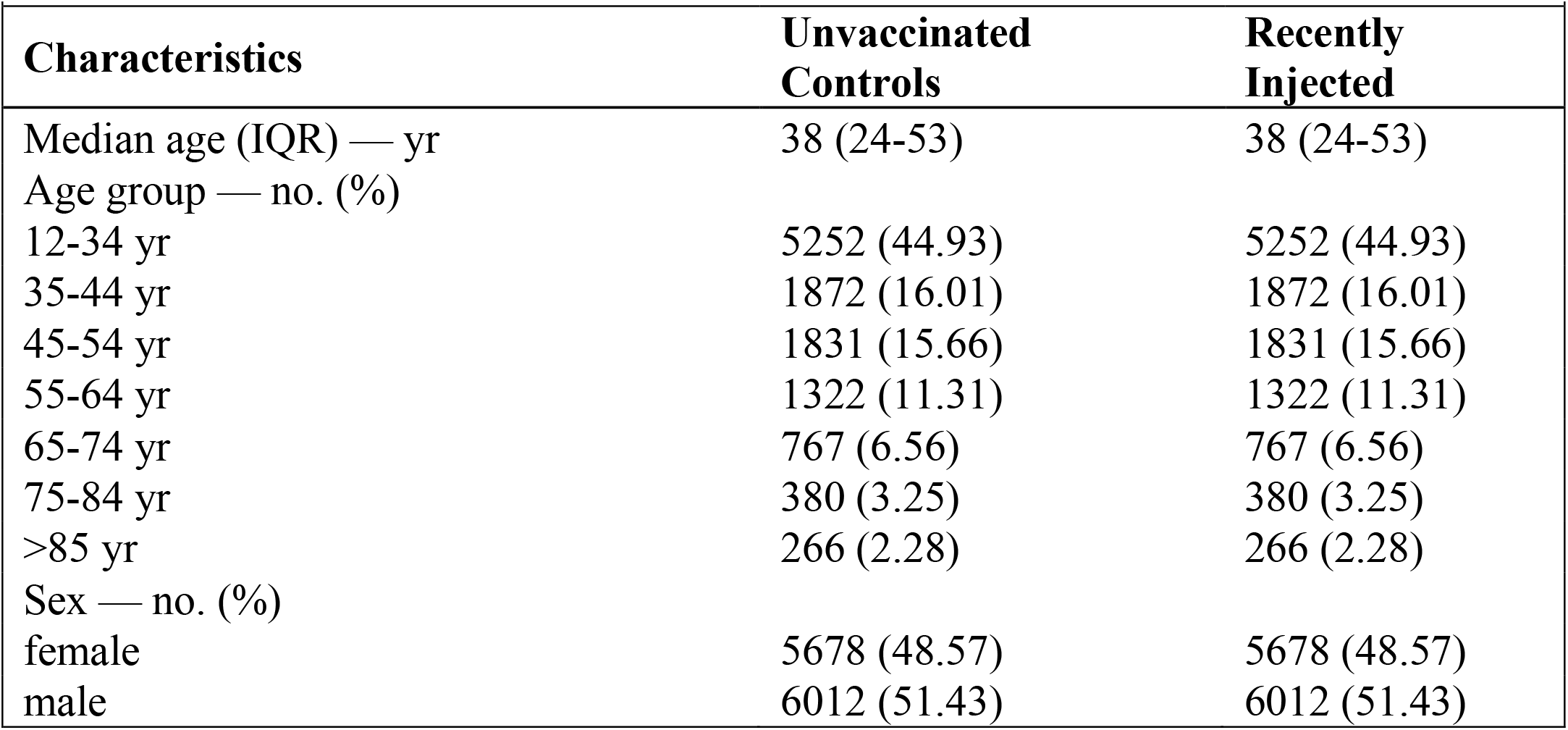
Demographic Characteristics of Recently Injected and Unvaccinated Control Groups.

### Primary Outcome

Mortality up to 60 days from first positive PCR, among infected people, 55 years of age and beyond, in all age groups and gender, was significantly lower in the recently injected group compared to the unvaccinated control group (Table 2). In the combined group of patients over 65 years and beyond, male and female, 143 deaths occurred among 1413 recently injected (10.12%) compared to 280 deaths among the 1413 unvaccinated (19.82%), odd ratio (OR) 0.46 (95% confidence interval (CI), 0.36 to 0.57; P<0.001) Fig. 2. Importantly, the most significant reduction in the death toll was observed among the combined 55 to 64 age group, with 8 deaths occurring among the 1322 recently injected (0.61%) compared to 43 deaths among the 1322 unvaccinated control (3.25%), OR 0.18 (95% CI, 0.07 to 0.39; P<0.001). Under the age of 55, we observed a very low death toll overall, with a borderline statistically significant difference between the groups. When combining all the patients aged 12-55 in both genders 10 deaths occurred among the 8955 recently injected group (0.11%) compared to 22 deaths among the 8955 unvaccinated control group (0.25%), OR 0.45 95% CI, 0.19 to 0.99; P=0.049). Analysis of each group separately did not yield significant difference, possibly due to the low death events in these age groups (Fig. S4 – Fig S9).

**Table 2.**
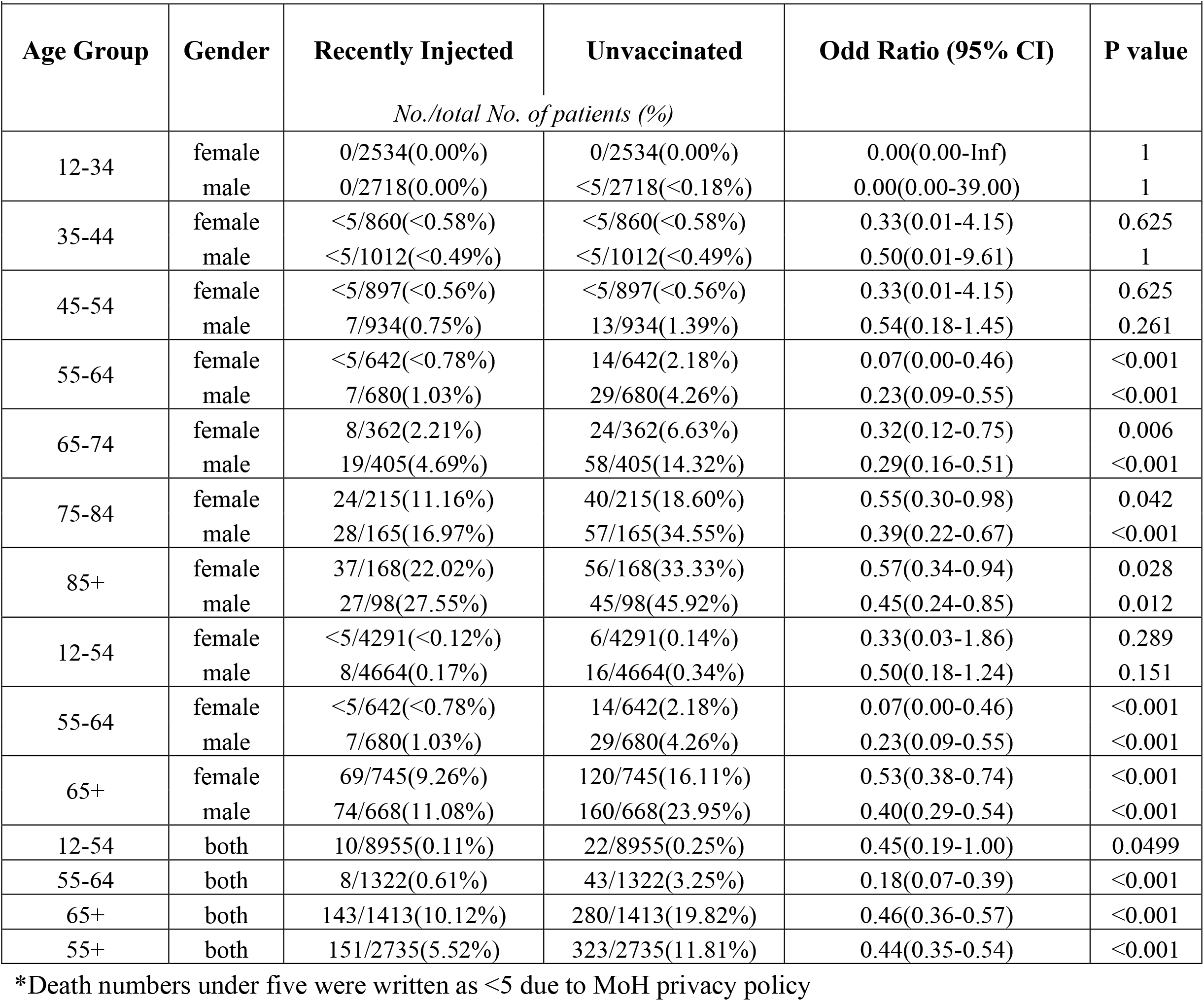
The Effect of Recent Vaccine injection on Mortality Rate in all age groups*.

**Figure 1.**
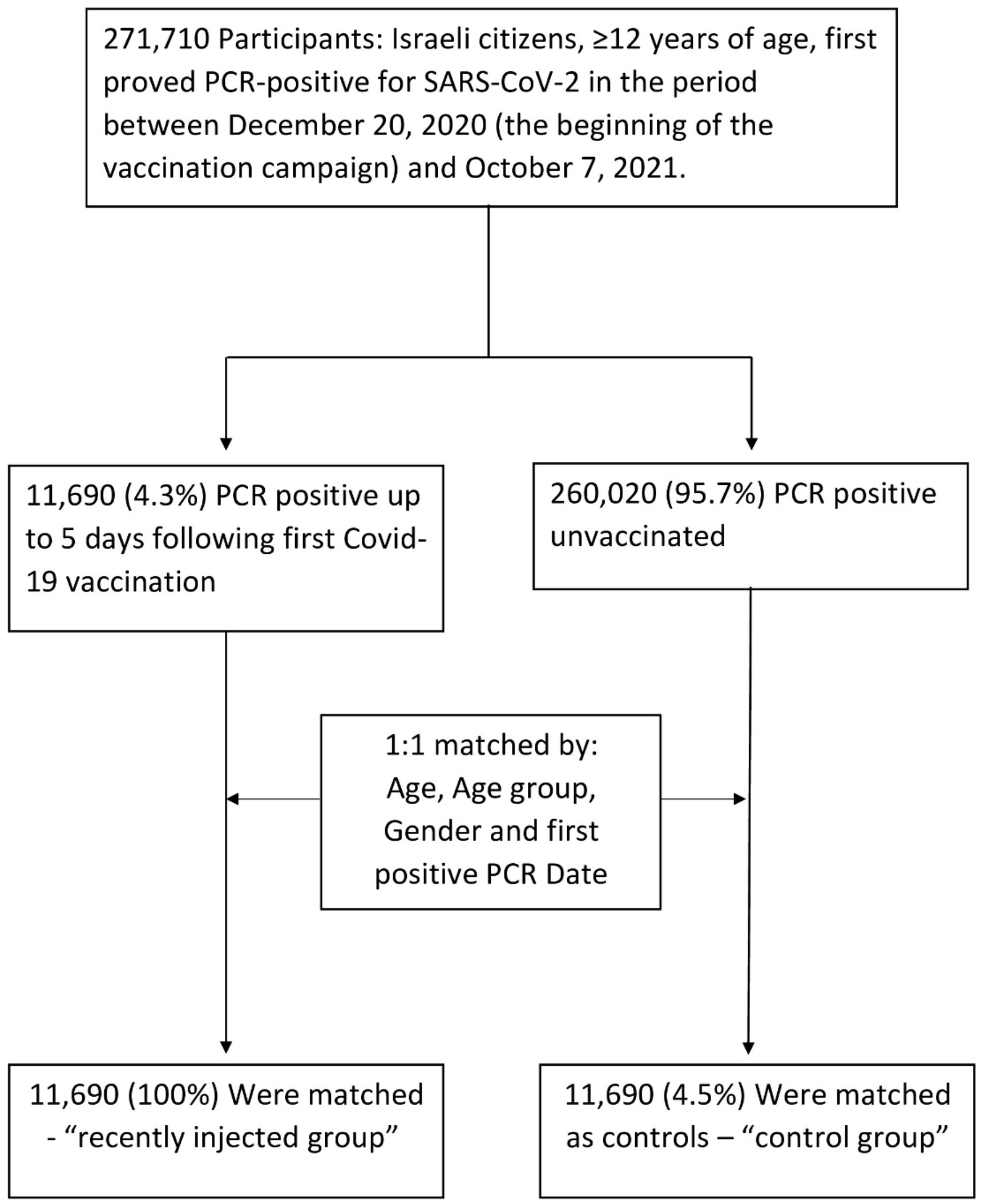
Study Population and Cohort Enrollment Process, December 20^th^, 2020, to October 7^th^, 2021. The 271,710 individuals that were included in the cohort, were all without previously documented SARS-CoV-2 PCR-positive results. Absolute numbers and percentage are shown for each group.

**Figure 2.**
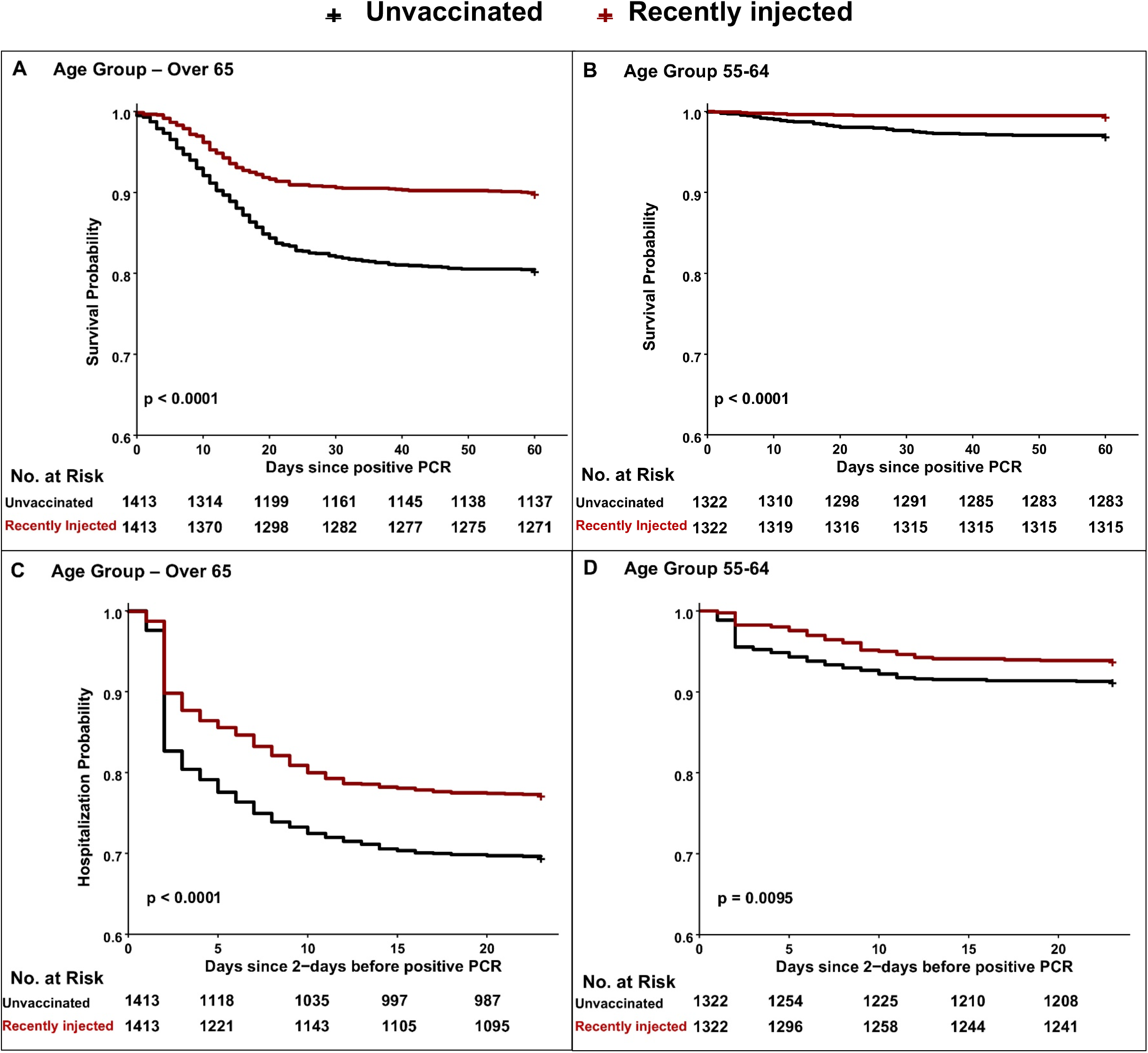
The clinical benefit of post-exposure-prophylaxis against SARS-CoV-2. Kaplan– Meier curves and the Number at Risk at each time point for mortality and hospitalization outcomes. **A**) Survival of age group 65 and over, starting from the day of the positive PCR. **B**) Survival of age group 55-64, starting from the day of the positive PCR. **C**) Hospitalization probability of age group 65 and over, starting from 2 days before the positive PCR. **D**) Hospitalization probability of age group 55-64, starting from 2 days before the positive PCR.

### Secondary Outcomes

Hospital admissions in the combined group of patients over 65 years and beyond, male and female, was significantly less in the recently injected group compared to unvaccinated group, 321/1413 (22.72%) versus 430/1413 (30.43%), respectively (OR 0.67 95% CI, 0.57 to 0.80; P< 0.001). A similar reduced hospitalization rate was seen in the 55-64 combined group, 81/1322 (6.13%) versus 115/1322 (8.70%), respectively (OR 0.69 95% CI, 0.50 to 0.93; P=0.014) (Fig 2, Table 3 and Fig S10-Fig. S15).

**Table 3.** The Effect of Recent Vaccine injection on Hospitalization Rate on all age groups.

| Age Group | Gender | Recently Injected<br>*No./total No. of patients (%)* | Unvaccinated | Odd Ratio (95% CI) | P value |
| --- | --- | --- | --- | --- | --- |
| 12-34 | female | 32/2534(1.26%) | 37/2534(1.46%) | 0.86(0.52-1.43) | 0.628 |
|  | male | 22/2718(0.81%) | 22/2718(0.81%) | 1.00(0.53-1.90) | 1 |
| 35-44 | female | 20/860(2.33%) | 23/860(2.67%) | 0.87(0.45-1.66) | 0.758 |
|  | male | 29/1012(2.87%) | 25/1012(2.47%) | 1.16(0.65-2.09) | 0.679 |
| 45-54 | female | 24/897(2.68%) | 35/897(3.90%) | 0.68(0.38-1.18) | 0.185 |
|  | male | 52/934(5.57%) | 40/934(4.28%) | 1.32(0.85-2.06) | 0.239 |
| 55-64 | female | 31/642(4.83%) | 36/642(5.61%) | 0.85(0.50-1.44) | 0.616 |
|  | male | 50/680(7.35%) | 79/680(11.62%) | 0.60(0.41-0.89) | 0.009 |
| 65-74 | female | 34/362(9.39%) | 49/362(13.54%) | 0.66(0.40-1.08) | 0.102 |
|  | male | 55/405(13.58%) | 102/405(25.19%) | 0.47(0.32-0.68) | <0.001 |
| 75-84 | female | 64/215(29.77%) | 76/215(35.35%) | 0.78(0.51-1.19) | 0.258 |
|  | male | 64/165(38.79%) | 72/165(43.64%) | 0.82(0.52-1.30) | 0.434 |
| 85+ | female | 59/168(35.12%) | 75/168(44.64%) | 0.67(0.42-1.07) | 0.095 |
|  | male | 45/98(45.92%) | 56/98(57.14%) | 0.64(0.35-1.16) | 0.153 |
| 12-54 | female | 76/4291(1.77%) | 95/4291(2.21%) | 0.80(0.58-1.09) | 0.164 |
|  | male | 103/4664(2.21%) | 87/4664(1.87%) | 1.19(0.88-1.60) | 0.272 |
| 55-64 | female | 31/642(4.83%) | 36/642(5.61%) | 0.85(0.50-1.44) | 0.616 |
|  | male | 50/680(7.35%) | 79/680(11.62%) | 0.60(0.41-0.89) | 0.009 |
| 65+ | female | 157/745(21.07%) | 200/745(26.85%) | 0.73(0.57-0.93) | 0.011 |
|  | male | 164/668(24.55%) | 230/668(34.43%) | 0.62(0.48-0.79) | <0.001 |
| 12-54 | both | 179/8955(2.00%) | 182/8955(2.03%) | 0.98 (0.79-1.22) | 0.915 |
| 55-64 | both | 81/1322(6.13%) | 115/1322(8.70%) | 0.69 (0.50-0.93) | 0.014 |
| 65+ | both | 321/1413(22.72%) | 430/1413(30.43%) | 0.67 (0.57-0.80) | <0.001 |
| 55+ | both | 402/2735(14.70%) | 545/2735(19.93%) | 0.69(0.60-0.80) | <0.001 |

Regarding the analysis on the length of hospitalization, we didn’t find any statistically significant effect of vaccination status. From our analysis, only age and gender play a role in the estimation of the hospitalization duration, with males and older age groups confirmed to increase the hospitalization length. No other interactions were significant.

## Discussion

The death toll from COVID-19 is rising, and numerous strategies are essential to reduce this mortality rate world-wide. In this report we show that the BNT162b2 vaccine could also be applied to patients as a post exposure prophylaxis manner, significantly reducing the death burden at the high-risk groups of age patients by 50%. We propose that this approach could be used with a significant positive impact on survival following SARS-CoV-2 exposure as well as upon hospitalization. There are numerous potential benefits in using this post-exposure prophylaxis approach. In many countries, once an individual is identified as positive by PCR for SARS-CoV-2, an epidemiological investigation is performed to identify those exposed to this COIVID-19 patient. These identified individuals are then isolated without any preventive treatment. We propose that as in other post exposure prophylaxis infectious events, the exposed individuals should be vaccinated to prevent severe disease culminating in hospitalization and in some cases leading to death. In many countries, the availability of vaccines is low. One question remains as to whom should the vaccine be provided. It is accepted in many societies that prioritization is to provide the vaccine to the aged, immunosuppressed patients and the medical staff. It could be also beneficial to include in this selected group of individuals, vaccination to SARS-CoV-2-exposed patients. This post-exposure prophylaxis could have an advantage of halting, or at least attenuating, the spread of the virus, possibly by reducing its titer in the respiratory track of exposed individuals. This approach is substantiated by the fact that infected patients are spreading the virus prior to the occurrence of symptoms (18).

Based on our findings, we propose that additional approaches should be investigated to potentially expand the usage of post exposure prophylaxis to reduce COVID-19 morbidity. Patients who develop severe respiratory symptoms, including low saturated oxygen (<93%), increased respiratory rate (>18/min) and pulmonary infiltrates on chest x-ray, could potentially benefit from a vaccination which would overcome the SARS-CoV-2 infection-related deterioration to pneumonitis and adult respiratory distress syndrome. Recently, an investigation that compared adult and children immune response to SARS-CoV-2 had shown that activation of the immune response in children results in an attenuation of infection (19). Stimulating the interferon response by vaccination could improve the immune reaction, simulating the children natural history of SARS-CoV-2 infection. It is well accepted that one important determinant in the outcome of COVID-19 infection is the genetic alterations in interferon pathway genes (20). Post-exposure prophylaxis could skew the immune response to enhance the interferon signaling pathways (21).

## Data Availability

Data are not available.

## Acknowledgment

The authors would like to thank Mrs. Nofar Avni, BI Developer from the IT Unit and Mrs. Olga Litinetsky (Slutsky) Head of Data Research Unit, Research Fund of the Hadassah Medical Organization for assistance in data accessibility and retrieval.

## Supplementary Materials

**Figure S1.**
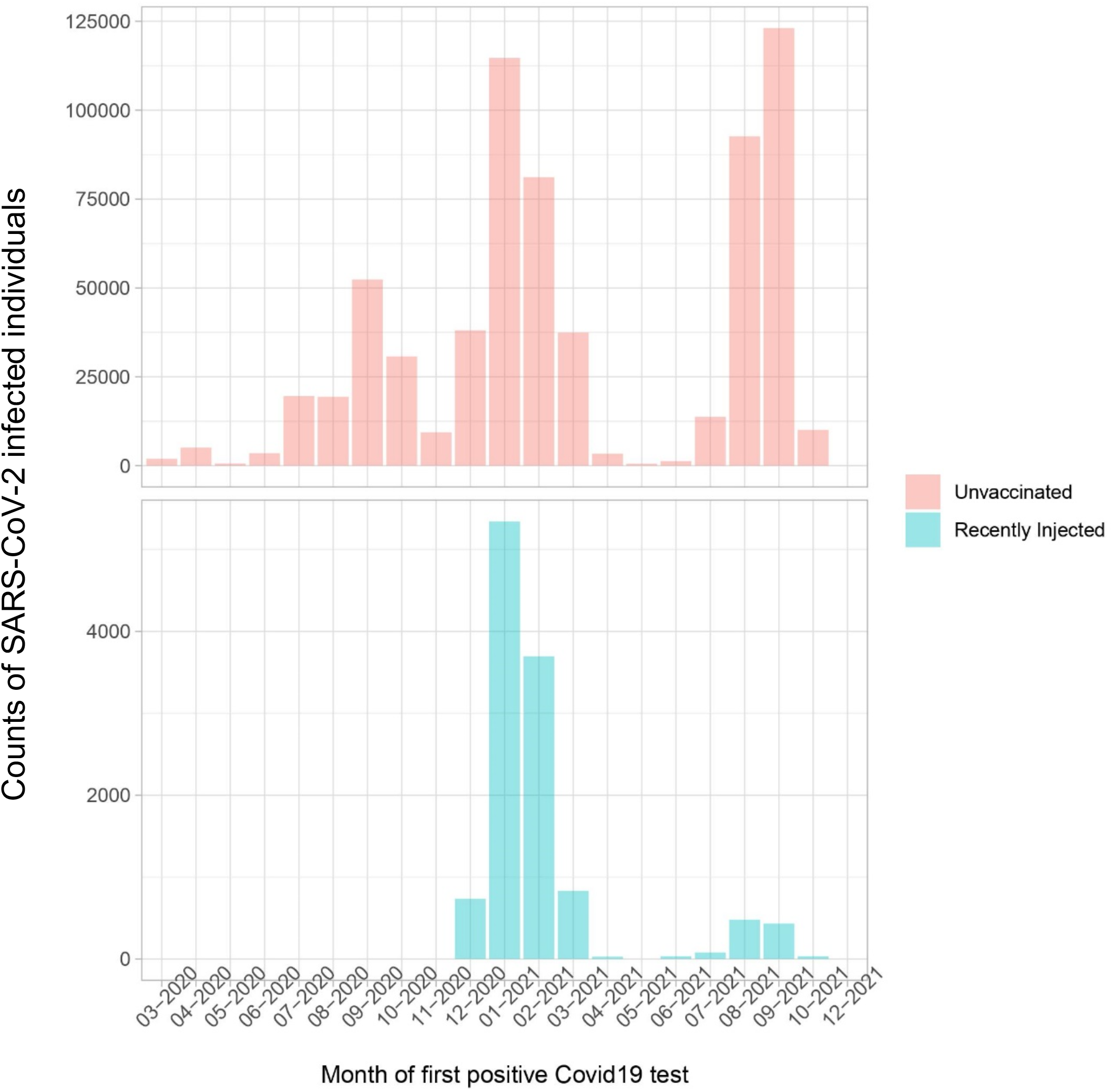
Monthly distribution of SARS-CoV-2 infected individuals. Absolute number of cases in the unvaccinated (upper panel) and the recently injected (lower panel) groups at each month since the beginning of the Covid-19 pandemic.

**Figure S2.**
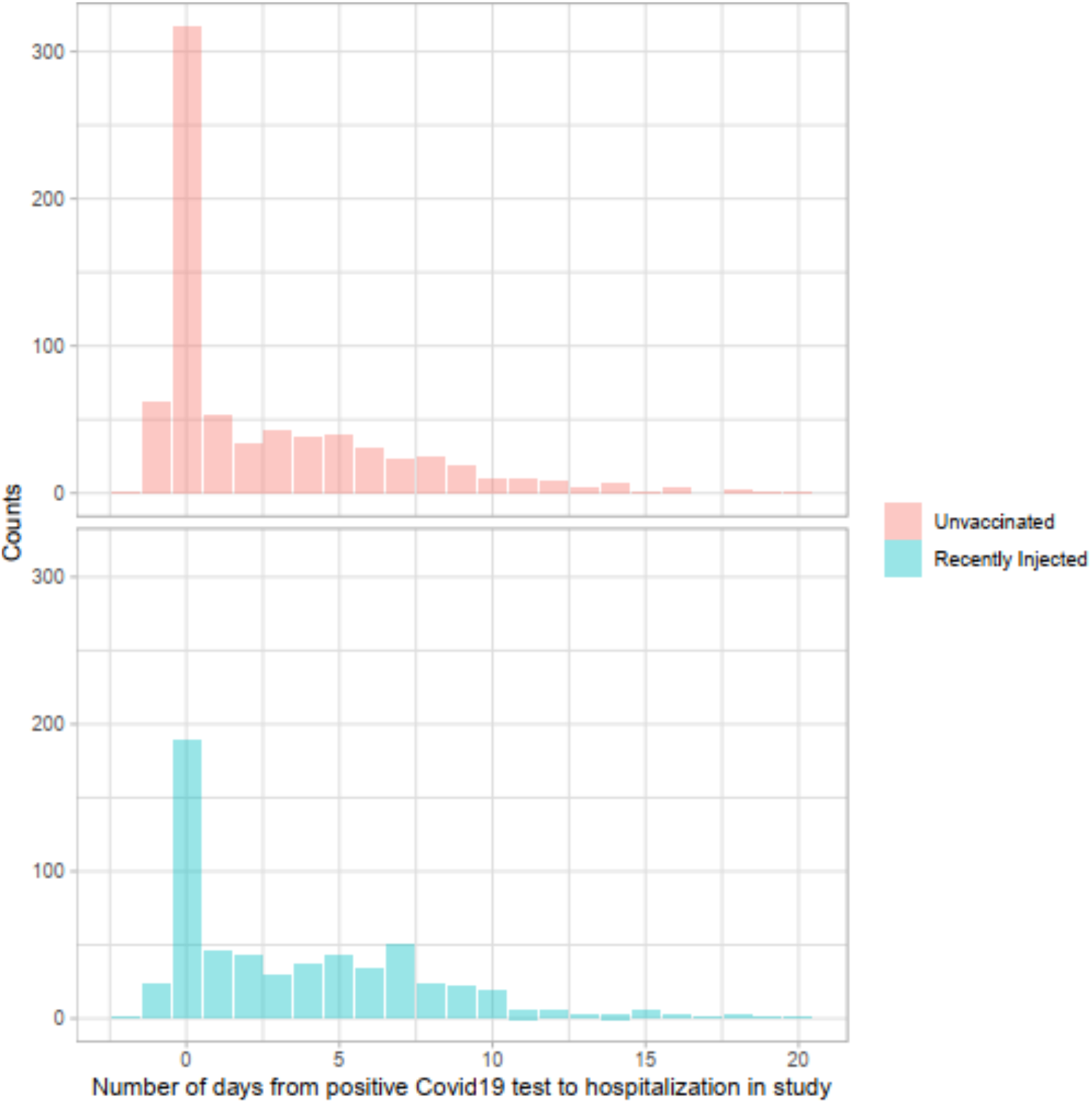
Time from Initial PCR Swabbing to hospitalization. Histograms showing the distribution of days number, from the date of the first positive PCR test until hospitalization arrival date. Unvaccinated (upper panel) and recently injected (lower panel).

**Figure S3.**
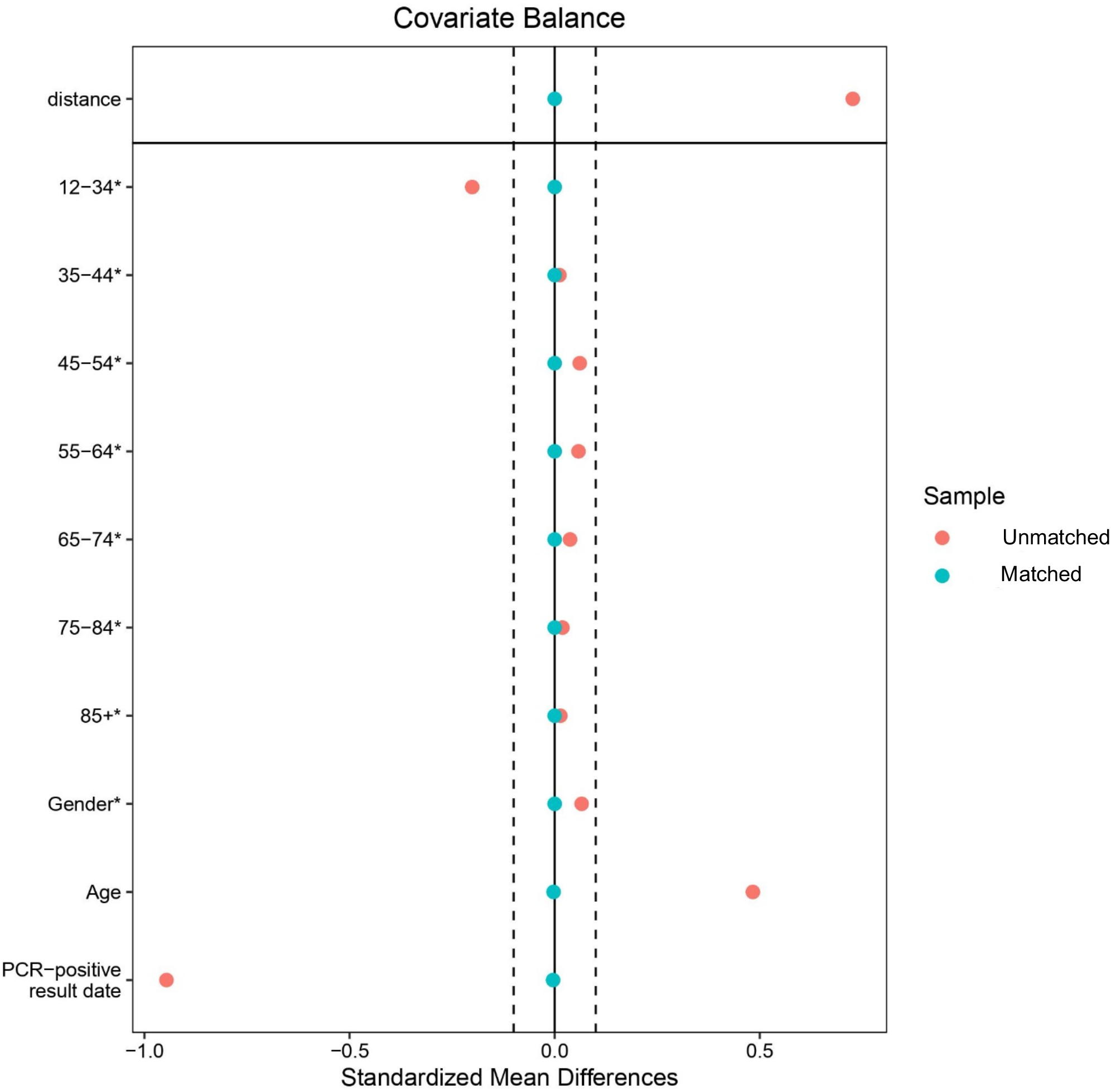
Covariate Balance (Love) Plot. A covariate balance (Love) plot showing the mean differences for continuous covariates and raw differences in proportion for binary covariates. A strict balance cut-off was set at 0.1.

**Figure S4.**
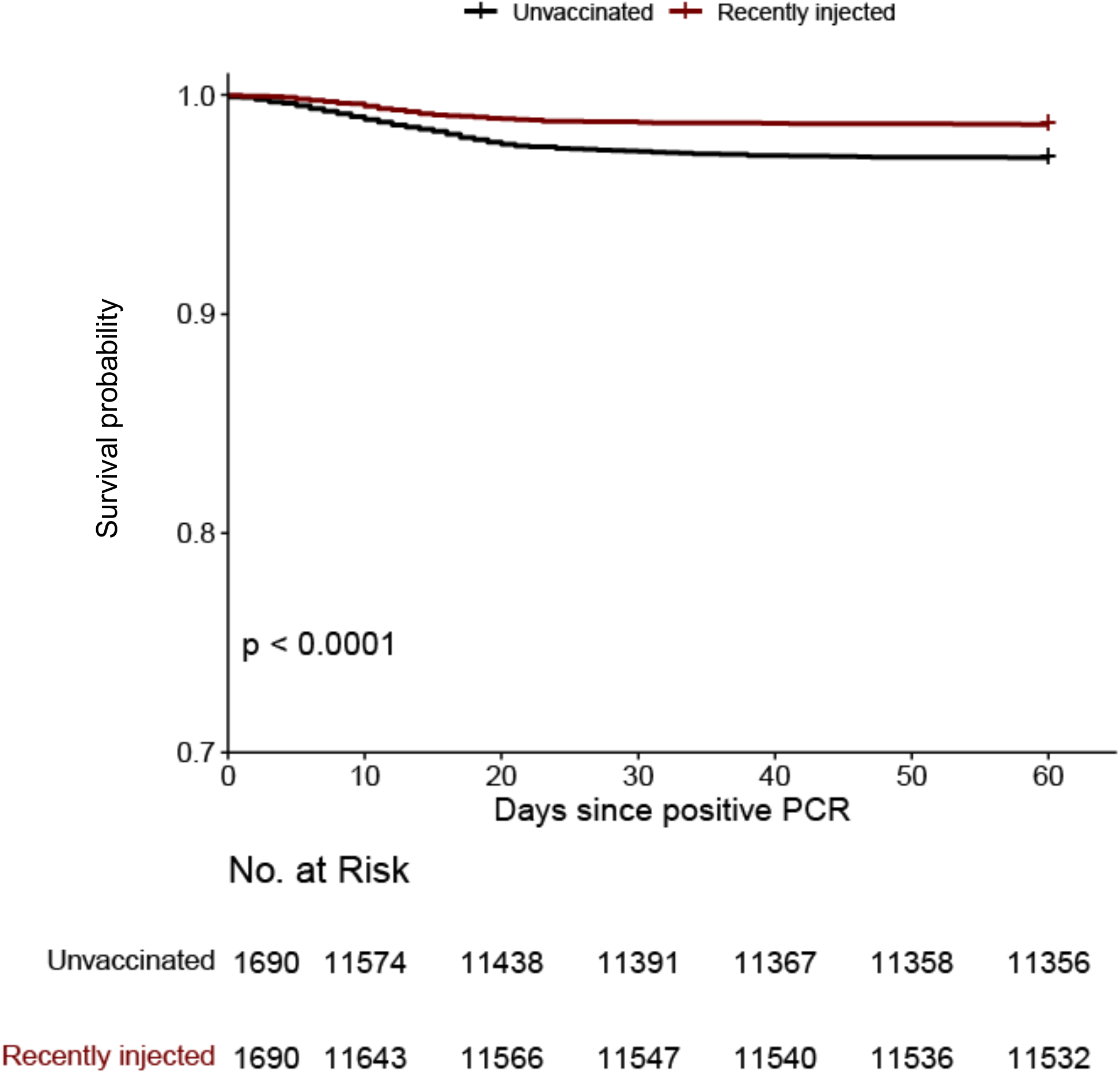
Survival probability of all patients in the study. Kaplan–Meier curves for survival and the Number at Risk at each time point of combined patients of each group (unvaccinated vs recently injected), starting from the day of the positive PCR.

**Figure S5.**
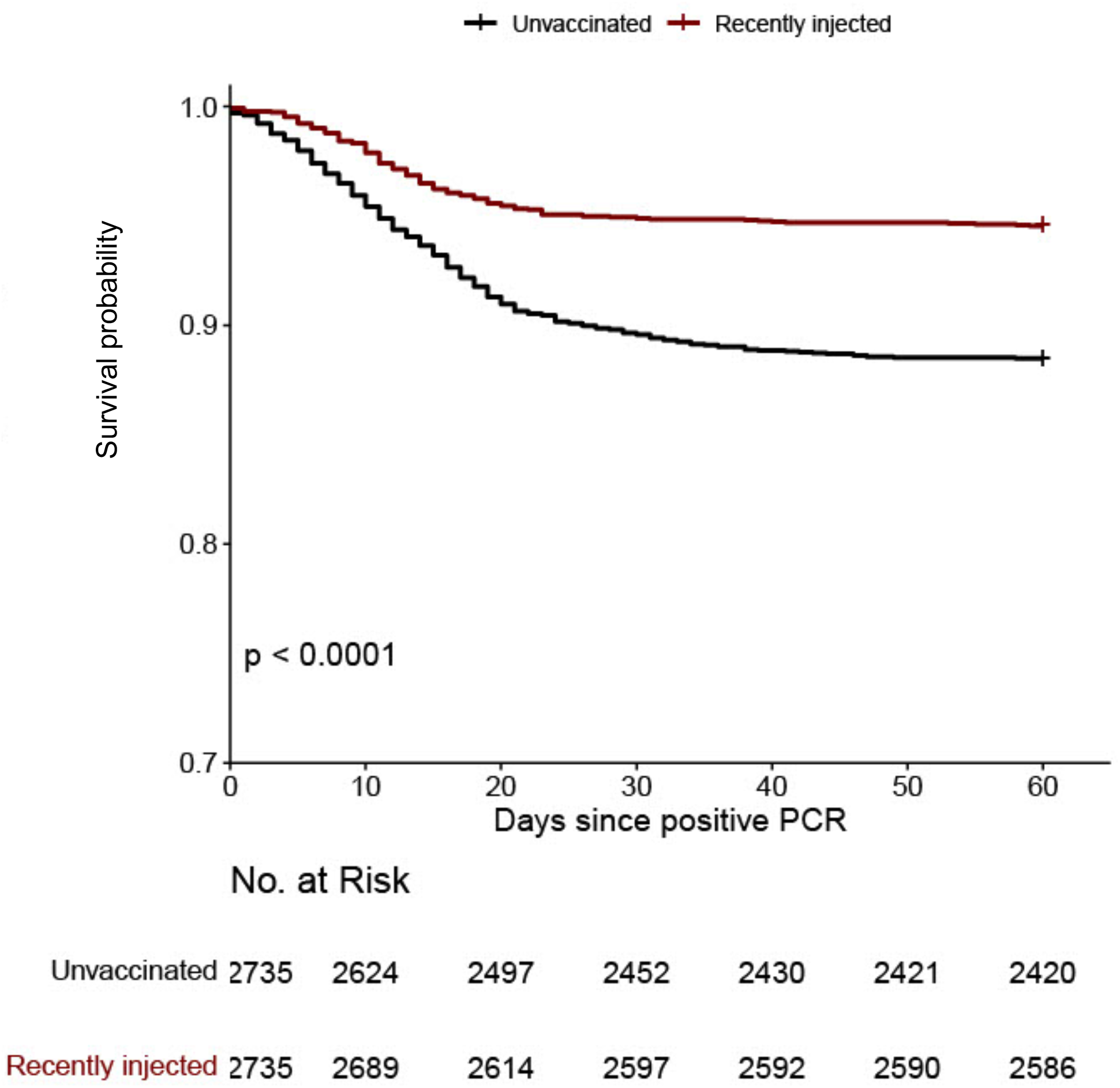
Survival probability of all patients 55 years and over. Kaplan–Meier curves for survival and the Number at Risk at each time point of age group 55 and over, starting from the day of the positive PCR.

**Figure S6.**
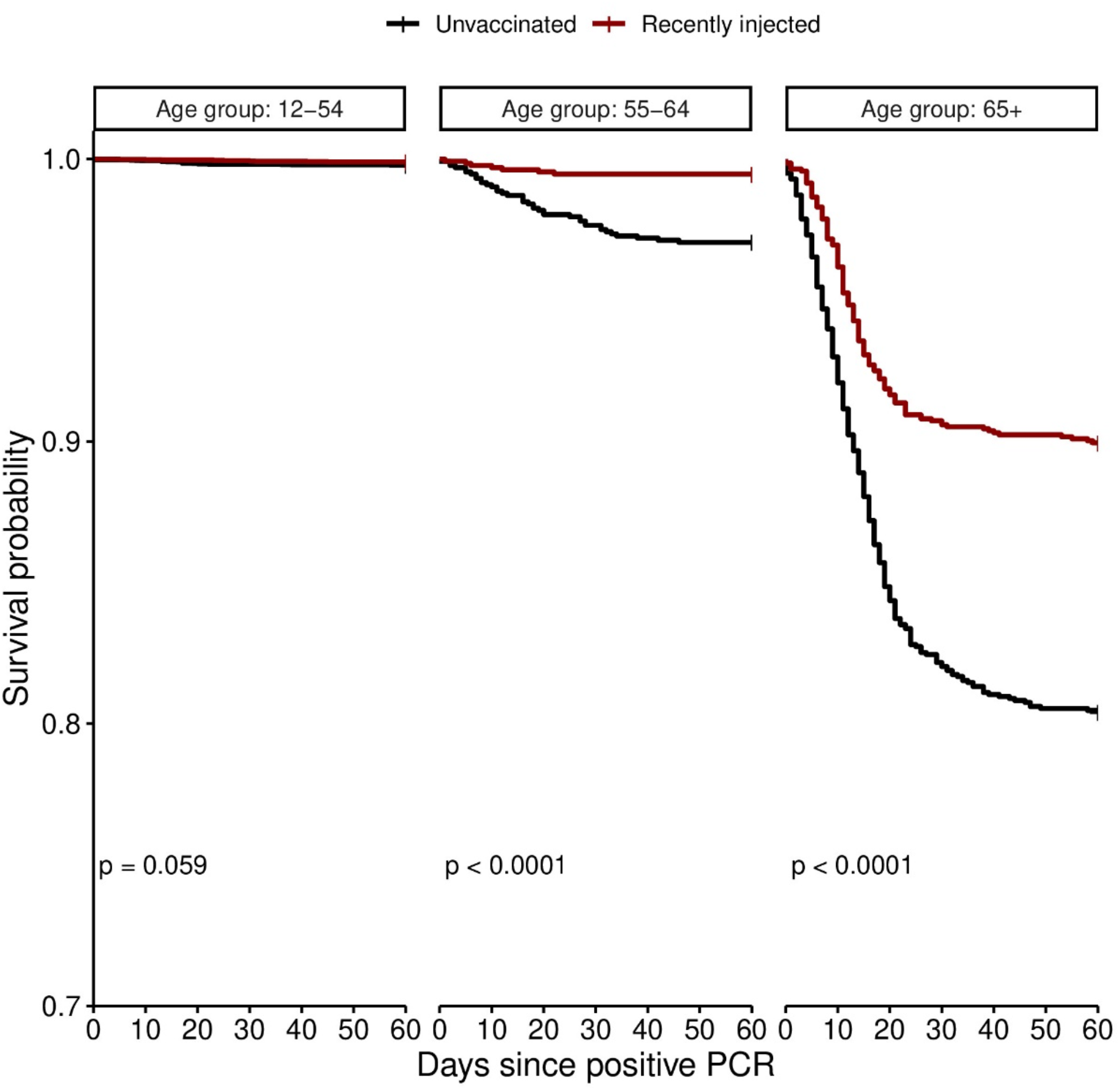
Survival probability of different age groups. Kaplan–Meier curves for survival of three combined age groups, starting from the day of the positive PCR.

**Figure S7.**
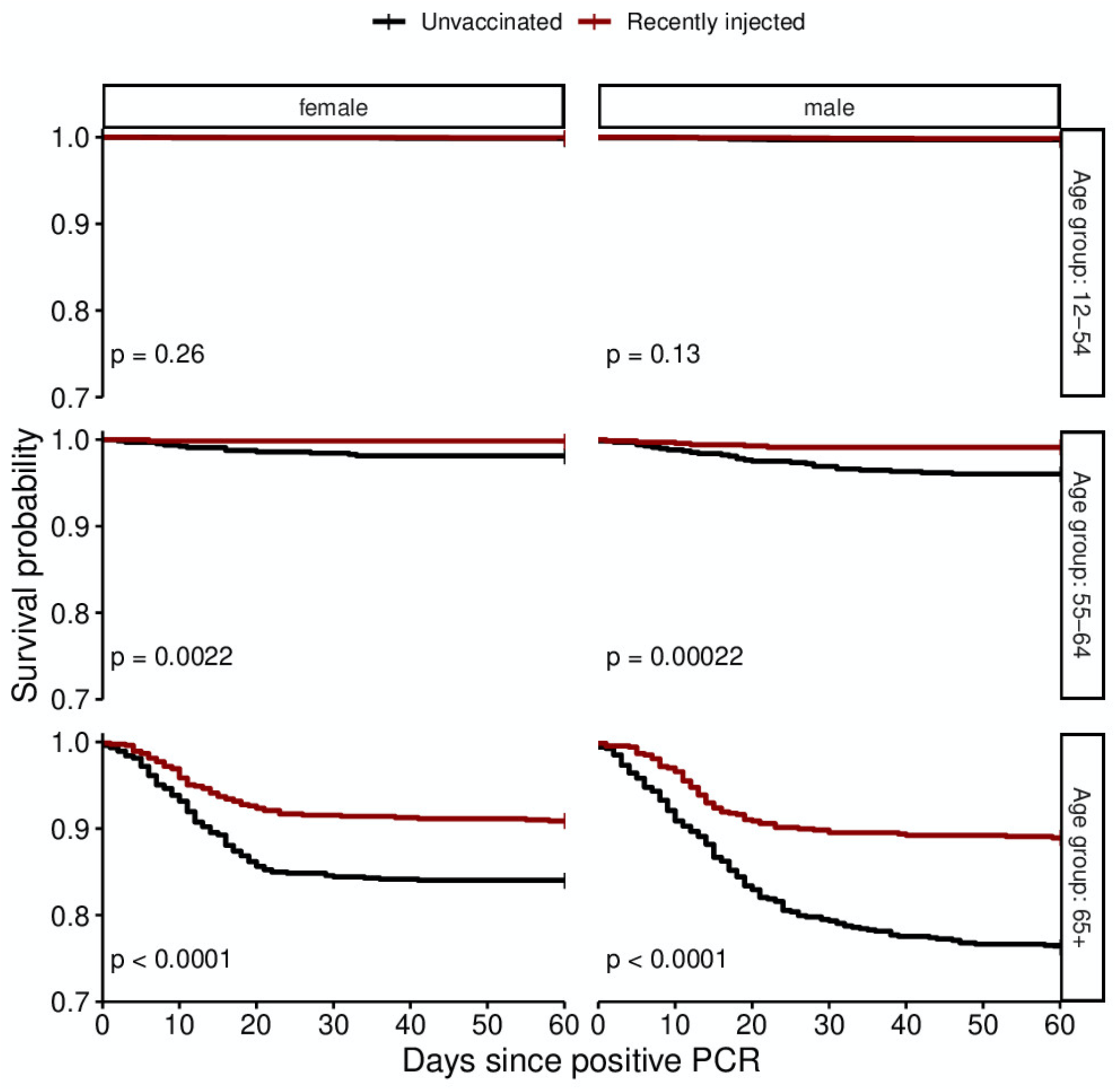
Survival probability of the different age groups divided by gender. Kaplan–Meier curves for survival of three combined age groups (shown in Fig S6) divided by gender, starting from the day of the positive PCR.

**Figure S8.**
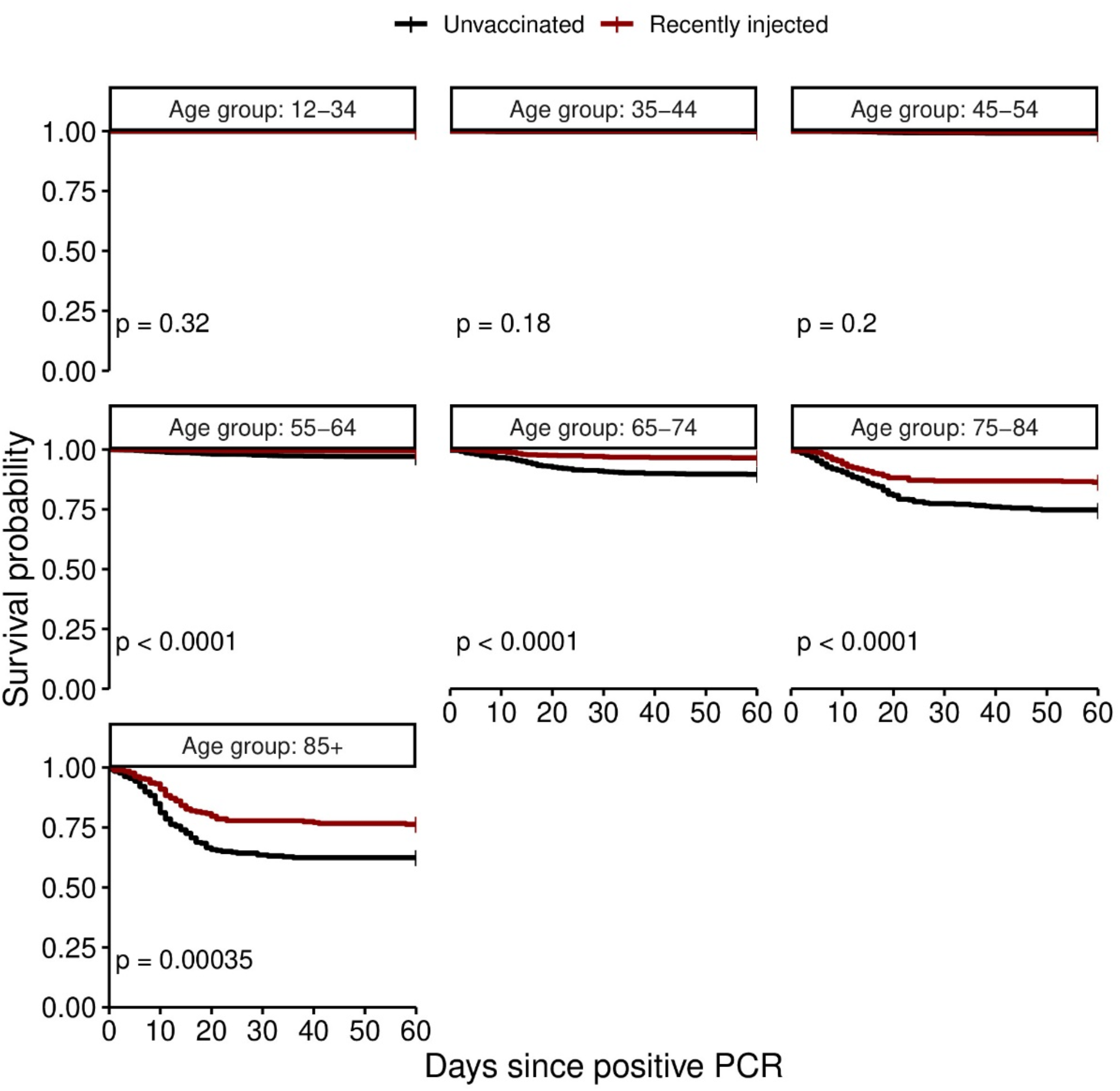
Survival probability of different age groups. Kaplan–Meier curves for survival of smaller distribution age groups (compared to age distribution in Fig. S6), starting from the day of the positive PCR.

**Figure S9.**
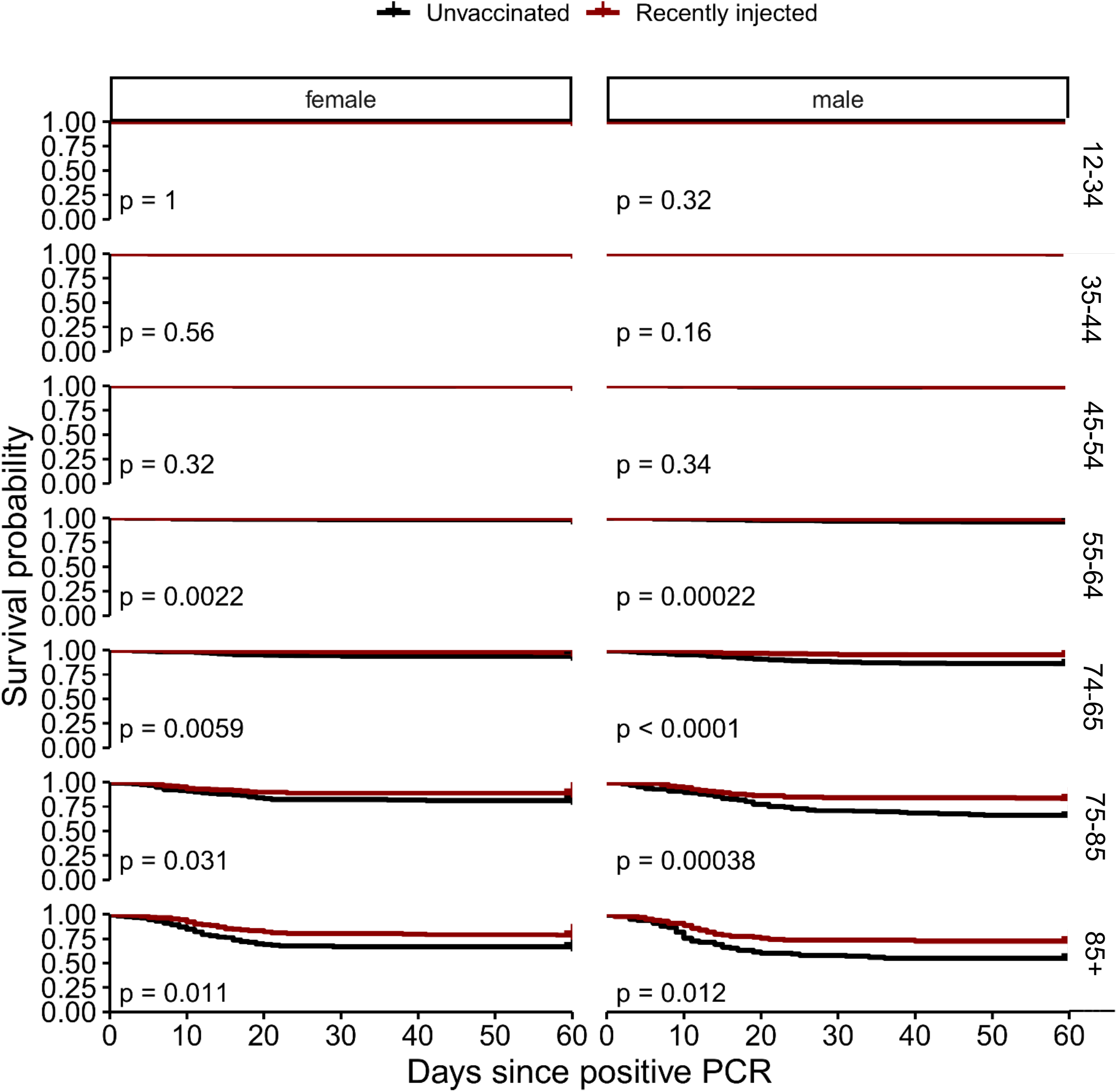
Survival probability of the different age groups divided by gender. Kaplan–Meier curves for survival of age groups shown in Fig S8, divided by gender, starting from the day of the positive PCR.

**Figure S10.**
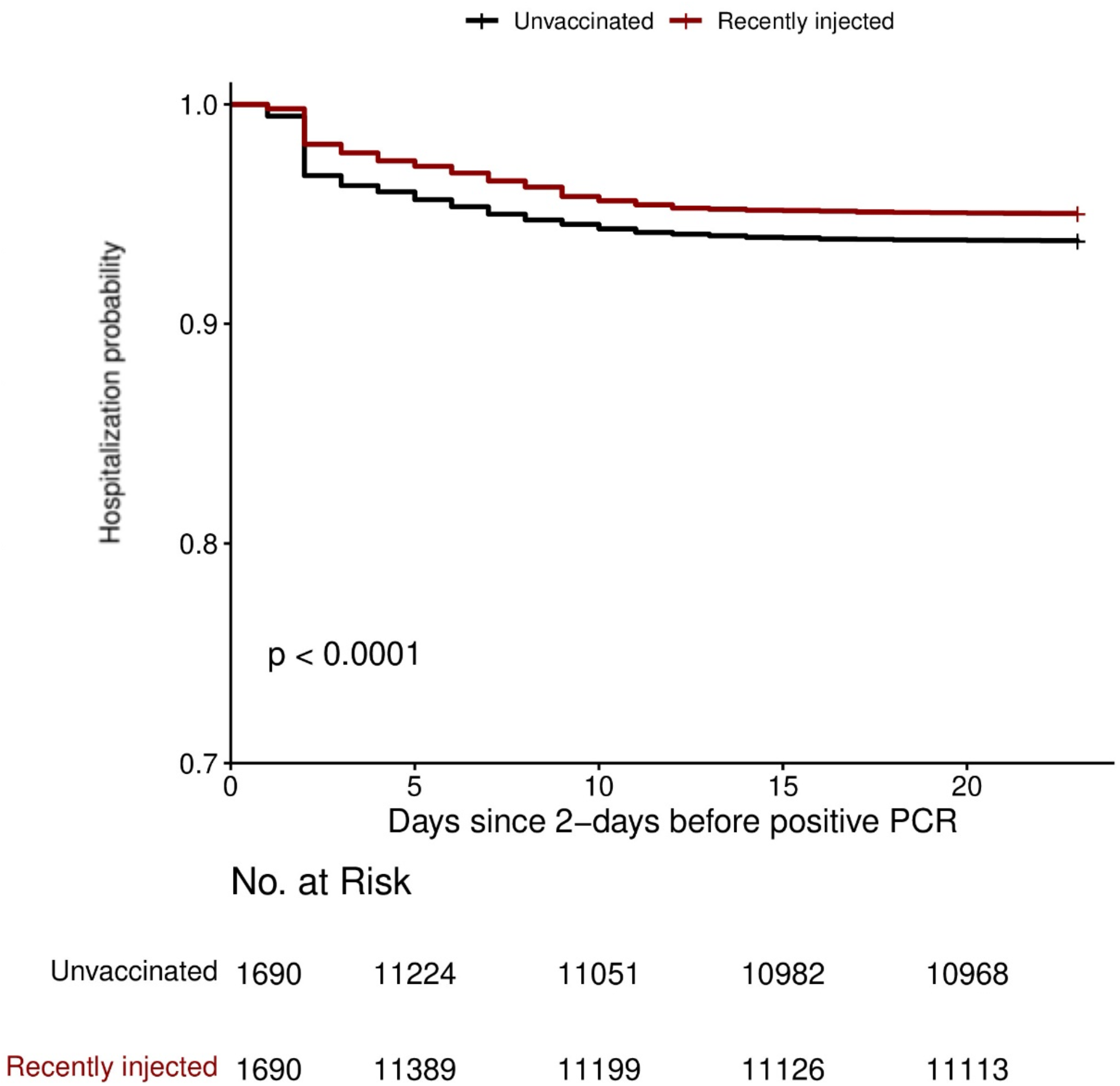
Hospitalization probability of all patients in the study. Kaplan–Meier curves for hospitalization and the Number at Risk at each time point of combined patients of each group (unvaccinated vs recently injected), Starting from 2 days before the positive PCR.

**Figure S11.**
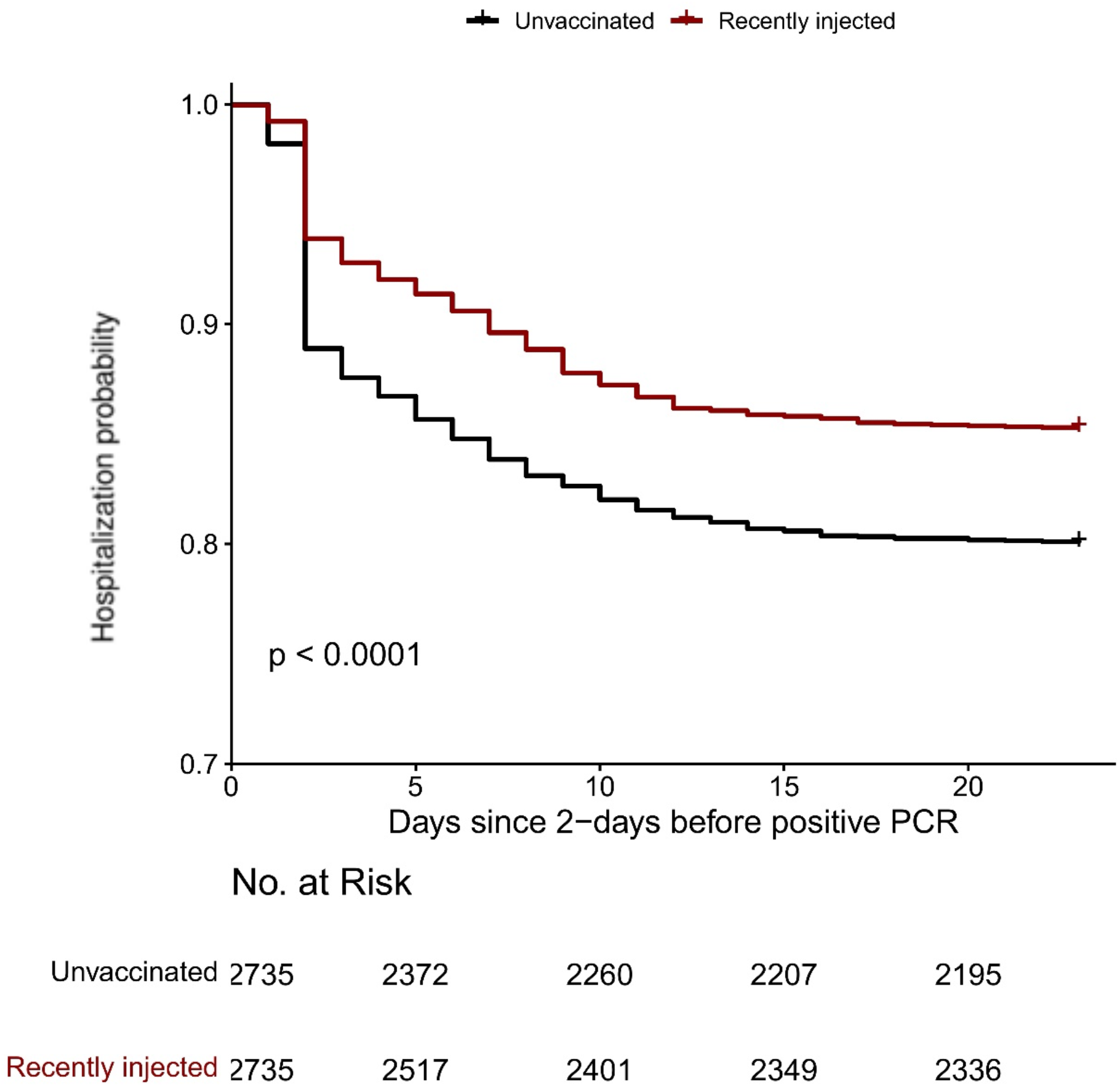
Hospitalization probability of patients 55 years and over. Kaplan–Meier curves for hospitalization and the Number at Risk at each time point of age group 55 and over, starting from 2 days before the positive PCR.

**Figure S12.**
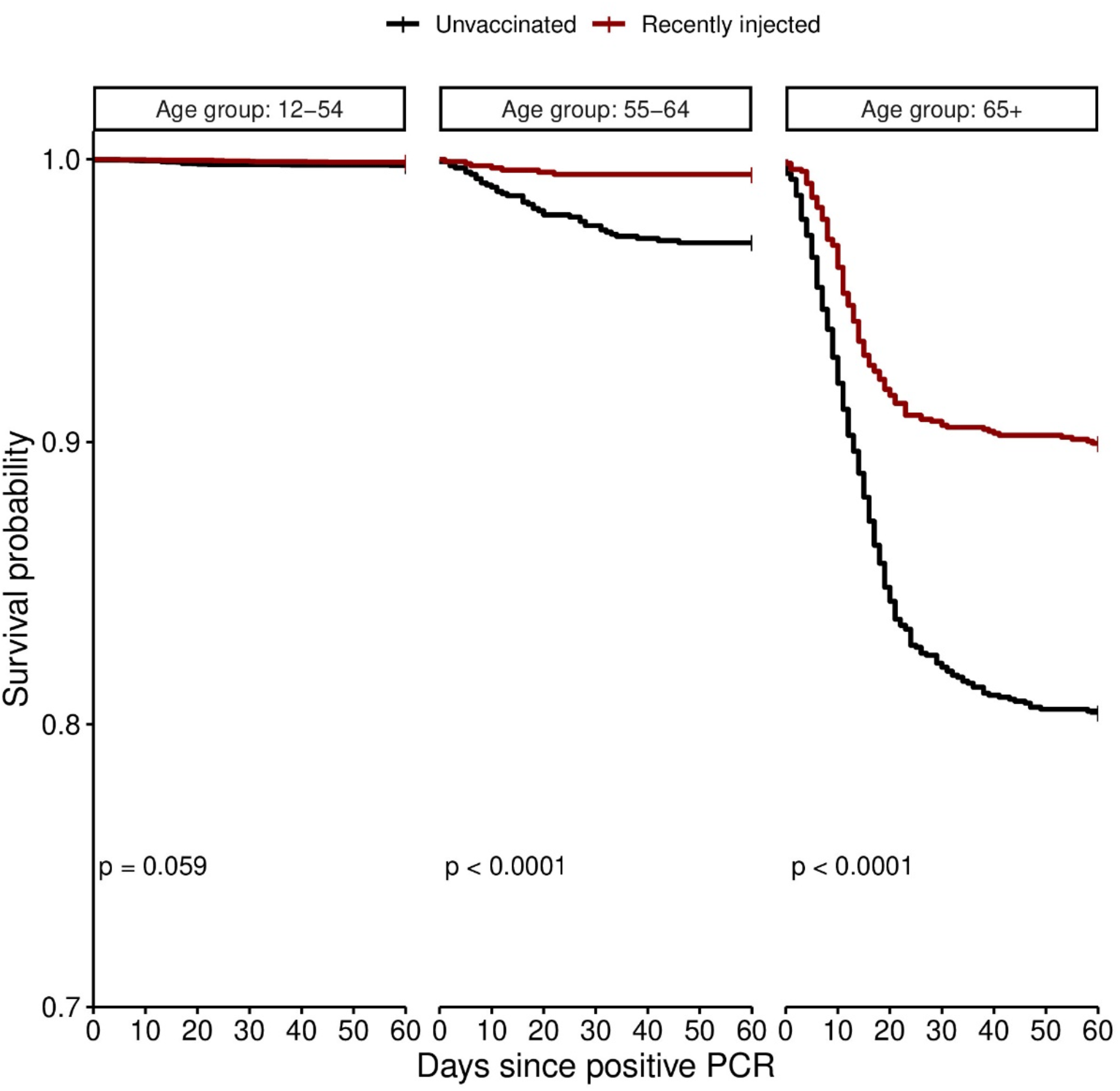
Hospitalization probability of different age groups. Kaplan–Meier curves for hospitalization of three combined age groups, starting from 2 days before the positive PCR.

**Figure S13.**
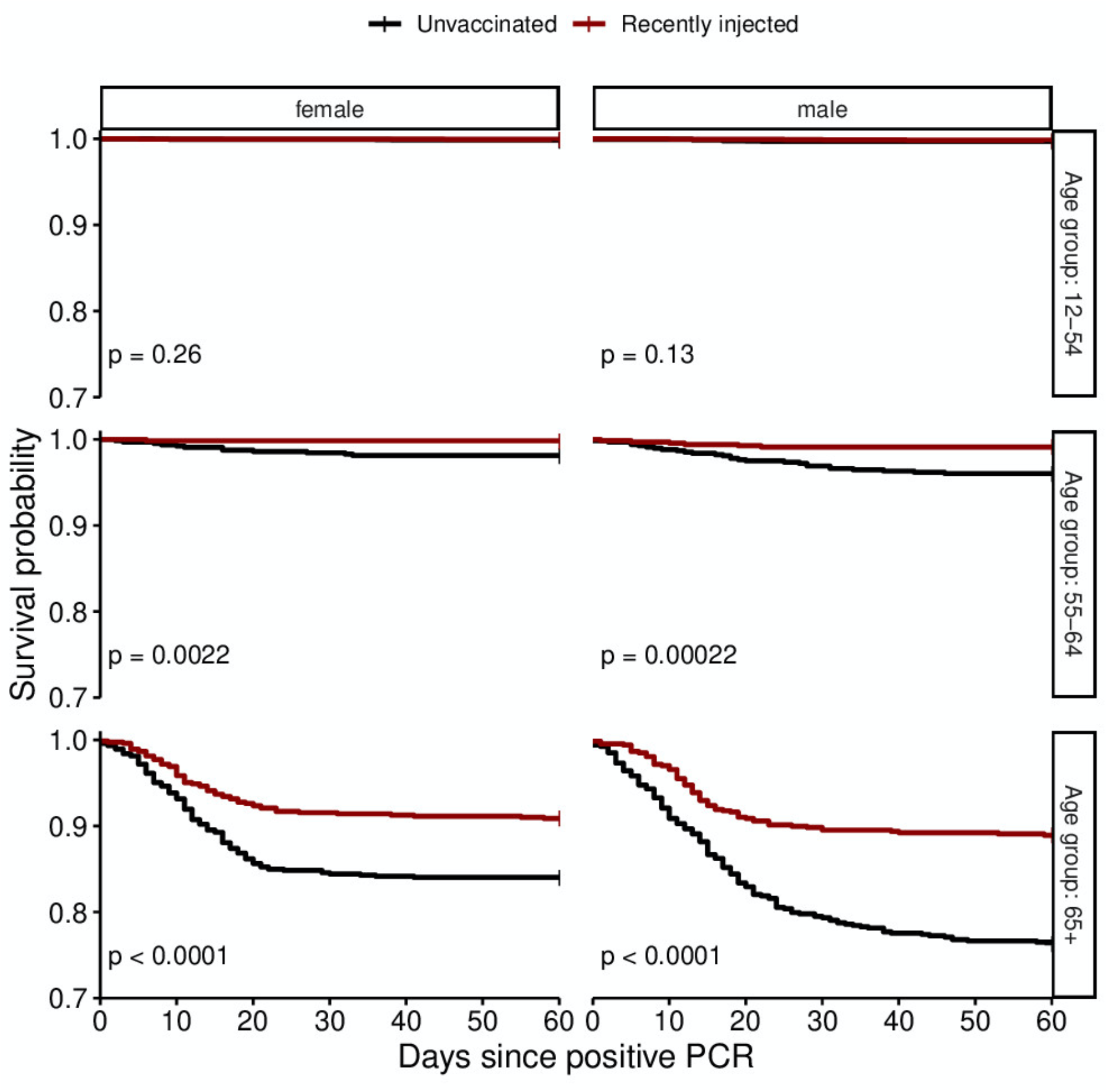
Hospitalization probability of the different age groups divided by gender. Kaplan–Meier curves for hospitalization of three combined age groups (shown in Fig S12) divided by gender, starting from 2 days before the positive PCR.

**Figure S14.**
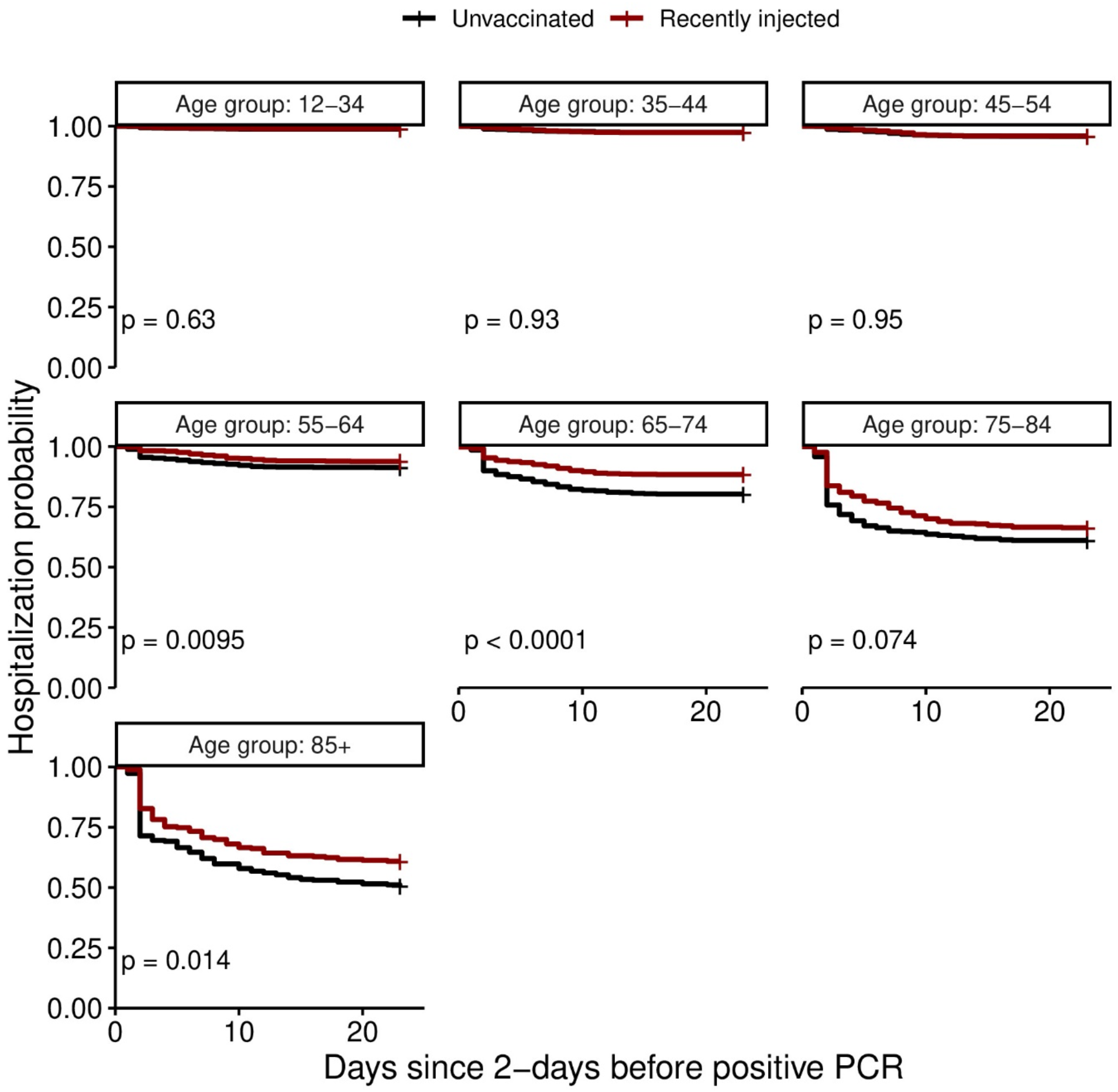
Hospitalization probability of different age groups. Kaplan–Meier curves for hospitalization of smaller distribution age groups (compared to age distribution in Fig. S12), starting from 2 days before the positive PCR.

**Figure S15.**
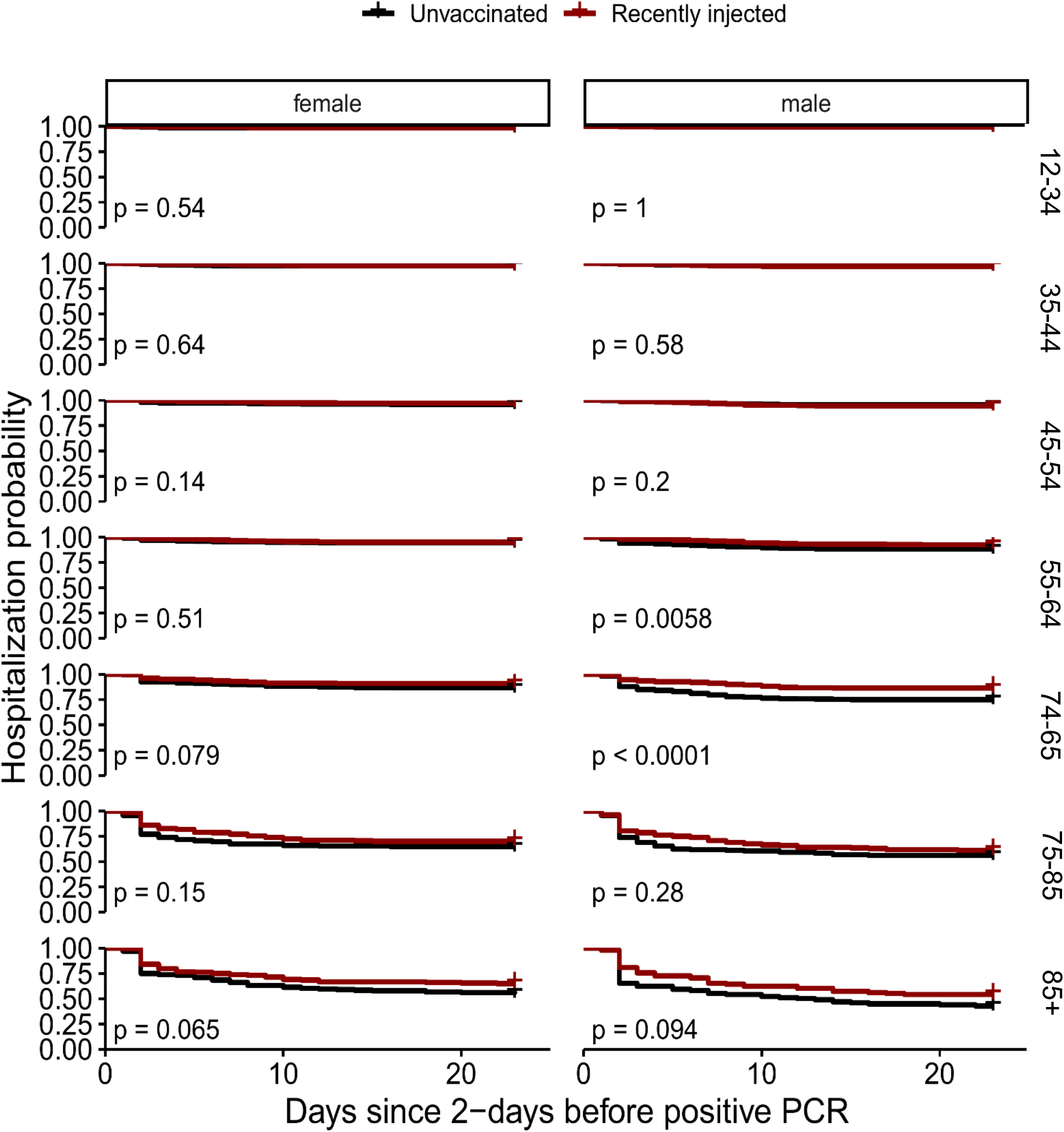
Hospitalization probability of the different age groups divided by gender. Kaplan–Meier curves for hospitalization of age groups shown in Fig S14, divided by gender, starting from 2 days before the positive PCR.

